# The gut microbiome associated with LGI1- and CASPR2-antibody encephalitis

**DOI:** 10.1101/2024.04.25.24305899

**Authors:** Edmund Gilbert, Sophie Binks, Valentina Damato, Christopher Uy, Paula Colmenero, Mohamed Ibrahim Khalil, Marcus O’Brien, Marcus Claesson, John F Cryan, Norman Delanty, Sarosh R Irani, Gianpiero L Cavalleri

**Affiliations:** School of Pharmacy and Biomolecular Sciences, Royal College of Surgeons in Ireland, Dublin, Ireland; FutureNeuro SFI Research Centre, Royal College of Surgeons in Ireland, Dublin, Ireland; Oxford Autoimmune Neurology Group, Nuffield Department of Clinical Neurosciences, University of Oxford, Level 3, West Wing, John Radcliffe Hospital, Oxford, OX3 9DS, UK; Dept of Neurology, John Radcliffe Hospital, Oxford, OX3 9DU; Department of Neurosciences Drugs and Child Health, University of Florence, Florence, Italy; Oxford Centre for Microbiome Studies, Kennedy Institute, University of Oxford, Oxford, OX3 7DQ, UK.SeqBiome Ltd., Moorepark Food Research Centre, Teagasc, Fermoy, P61 C996 Cork, Ireland; APC Microbiome Ireland, University College Cork, T12 YT20 Cork, Ireland; School of Microbiology, University College Cork, T12 YT20 Cork, Ireland; Department of Anatomy and Neuroscience, University College Cork, T12 YT20 Cork, Ireland; Departments of Neurology and Neurosciences, Mayo Clinic, Jacksonville, Florida, USA

## Abstract

Autoimmune encephalitis is a cause of brain inflammation characterised by auto-antibodies which target cell surface neuronal proteins, and lead to neuronal dysfunction. In older people, common forms are encephalitis with autoantibodies to leucine-rich glioma inactivated protein 1 (*LGI1*) and contactin associated protein like 2 (*CASPR2*), whose presentation includes frequent focal seizures. The exact cause of these autoantibodies remain unknown, but established predispositions include overrepresented human leukocyte antigen (HLA) alleles. Yet, these alleles are themselves common in the healthy ancestry-matched population. One potential aetiological hypothesis is that an environmental trigger, such as the gut microbiome, interacts with a genetically predisposed individual. To investigate this, we studied 47 patients with leucine-rich glioma-inactivated 1 (LGI1)- or contactin-associated protein 2 (CAPSR2)-antibody encephalitis (LGI1/CASPR2-Ab-E) and 37 familial/environmentally matched controls, and performed metagenomic shotgun sequencing, to describe compositional and functional differences in the gut microbiome. We observed that LGI1/CASPR2-Ab-E gut microbiomes exhibited a significant reduction in the ratio of *Firmicutes* and *Bacteroidetes* phyla, which associated with dosage of HLA susceptibility alleles in LGI1-Ab-E patients. Furthermore, we identified differences in functional gene profiles in the gut microbiome that led to a reduction of neuroinflammatory protective short-chain-fatty-acids (SCFA) in LGI1-Ab-E patients. Taken together, our results suggest that a compositional shift in the gut microbiome of LGI1/CASPR2-Ab-E associates with a neuroinflammatory state, possibly through the reduction of SCFA production. Our study highlights the potential of the gut microbiome to explain some of the complex condition and unravel aetiological questions. Validation studies with greater sample sizes are recommended.

## Introduction

Autoimmune encephalitis (AE) is characterised by the presence of autoantibodies with pathogenic potential which target cell surface neuronal proteins^1–3^. Among the most common autoantibodies detected in AE are those which bind leucine-rich glioma inactivated protein 1 (LGI1) and contactin associated protein like 2 (CASPR2). Many of these patients typically have very frequent, focal seizures in addition to altered mood, personality change, and cognitive impairment^4^. This semiology of these seizures can be pathognomonic, as is well-established for patients with faciobrachial dystonic seizures in association with LGI1-antibodies^5–7^. In addition, other unusual semiologies in LGI1-antibody encephalitis (LGI1-Ab-E) include thermal, piloerection, autonomi and emotional events^8,9^. Patients with CASPR2-antibodies also have very frequent focal seizures ^10–12^. These clinical observations imply that autoantibodies mediate forms of seizures which are likely to have characteristic, and sometimes novel, underlying molecular mechanisms.

In addition to distinctive clinical features, these AE patients are often anti-seizure medication (ASM) resistant, but show beneficial responses to immunotherapies, including corticosteroids, plasma exchange, and intravenous immunoglobulins^5,6,13,14^. Yet, despite immunotherapies and ASMs, more than 70% of patients are left with long term deficits, which impair quality of life^5,15,16^. In addition, around 40% of people treated with immunotherapies experience adverse drug effects^5,6^, and in the case of LGI1-Ab-E up to 20% develop chronic epilepsy^17^. Hence there is an unmet need for personalised immunotherapies that could be safe and effective in the long term.

Despite its potential to explain causation and address therapeutic needs, the aetiology of these diseases is poorly understood. Recent works have demonstrated that more than 90% of patients with LGI1-antibodies carry specific class-II HLA alleles, namely HLA-DRB1*07:01 (OR: 26.4 [CI=8.5-81.5]^18^, OR: 85.5 [CI=10.4-699.6]^19^). Whereas CAPSR2-antibody AE show a strong but distinct HLA profile with an over-representation of HLA-DRB1*11:01^20^ (OR: 9.4 [CI=4.6-19.3]^21^). A striking feature of these genetic findings is the relatively common underlying population frequencies of the HLA risk alleles. The HLA-DRB1*07:01 allele, for example, is also carried by 27% of healthy European-ancestry populations^18,21^ (∼12% globally^22^) and HLA-DRB1*11:01 is present in 9% of the general European-ancestry population^21^ (∼5% globally, maximal in Europe^22^). Whilst these strong HLA Class II associations clearly implicate T cells in disease pathogenesis, their high general population frequencies only partially explain disease causation. Hence, we examined potential contributory environmental triggers.

The gut microbiome elegantly provides an interface to explain such an environmental trigger. Indeed, in a variety of autoimmune central nervous system diseases, there is growing evidence for an aetiological link between the gut microbiome and the brain, principally via innate immunity and T-cells^23–25^. In several autoimmune diseases, skews in gut microbial populations have been discovered which may predispose to the illness^23,26–28^. Further, in autoantigen-specific illnesses, a distinctive microbial signature may provide a similarly elegant link to the LGI1 and CASPR2 autoantigens through molecular mimicry.

Here, we investigated the gut microbiome in patients with LGI1- and CASPR2-Ab-E. We also collected stool and salivary DNA from matched but unaffected relatives, close family members, or friends. We sought to; 1) determine if species abundance or diversity in the gut microbiome was significantly different between healthy and affected family members, 2) characterise potential functional differences in these microbiotas, and finally 3) investigate the evidence of LGI1 sequence homologues within the gut microbiota of LGI1-Ab-E patients.

## Results

### Recruitment and Cohort

We recruited 47 patients with LGI1-(n=43), CASPR2-antibody (n=3), or double positive LGI1/CASPR2-antibody (n=1) encephalitis, henceforth collectively referred to as LGI1/-CASPR2-Ab-E. In parallel, we recruited matched healthy controls (HC, n=37) through close family members who were typically a domestic partner (see **Table 1**). We isolated both genomic DNA from blood and gut microbiome DNA from stool (see Methods). All cases and HC were consistent with British- or Irish-like ancestry in comparison to ancestry controls (**Supp Figure 1**). Tissue typing confirmed an enrichment of the HLA DRB*07:01 allelotype in LGI1 patients (**Table 1**) compared to SNP genotype imputation of HLA alleles in HC.

**Figure 1.**
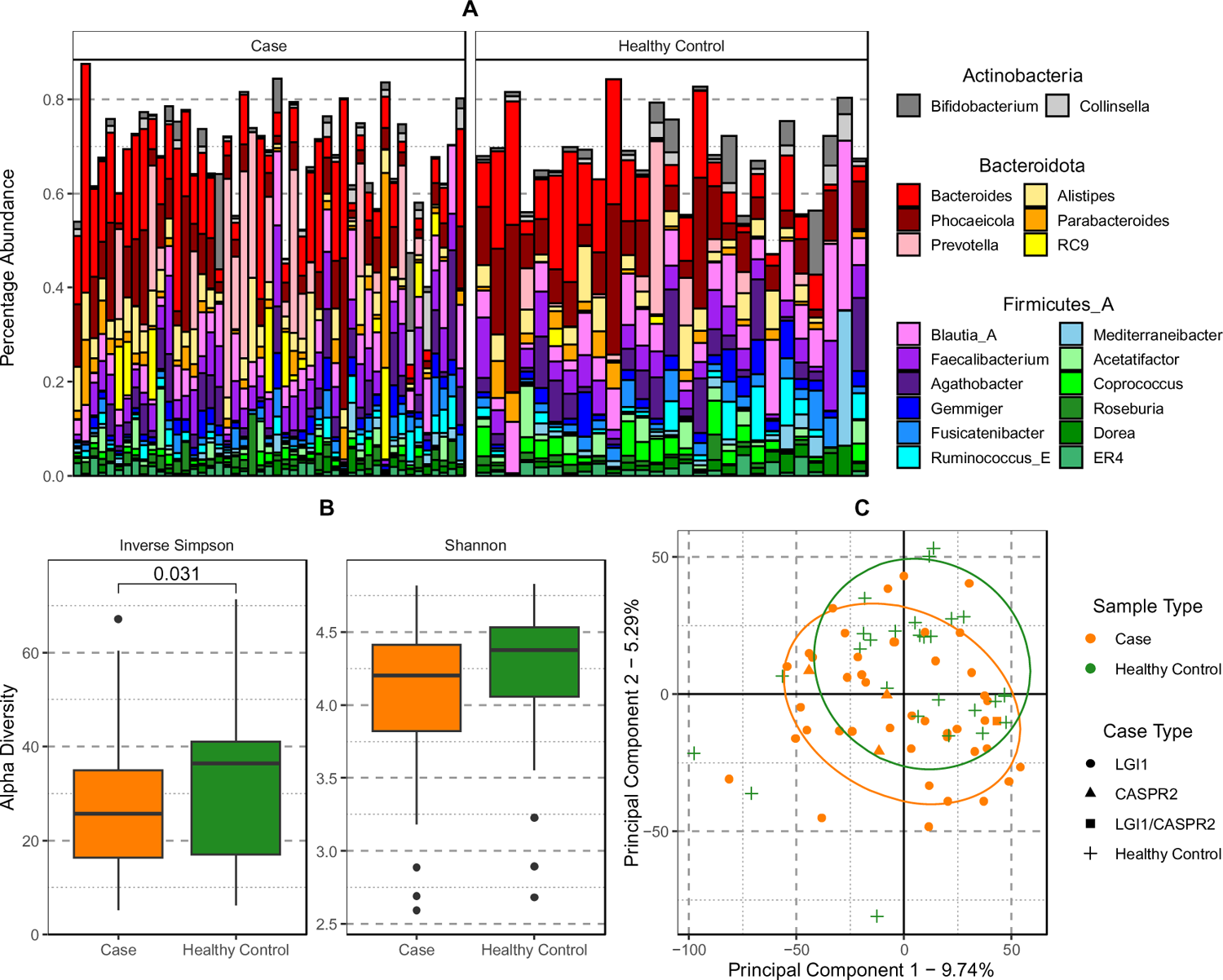
Comparison of the gut microbiome in *LGI1*-*CASPR2*-Ab-E cases and healthy controls. – (A) Bar chart of the 20 most abundant Genera, grouped by Phylum and separating cases (left) and healthy control (HC) samples (right) in separate panels. Each vertical bar represents the compositional microbiome profile of an individual. Note: Firmicutes_A and similar names are placeholder taxa which have been classified as such in the NCBI database but does not meet the clustering requirements for GTDB. Unused space represents other species not in the top 20 most abundant. (B) The distribution of alpha diversity values between cases (orange) and HC (green) using two measures, the Inverse Simpson and Shannon to retain comparative direction of effect between the two. Pairwise p-values shown are Wilcoxon test p-values significant after Holm correction for multiple testing (Shannon not significant). (C) Principal Component Analysis of beta-diversity estimates between cases and HC. Ellipses shown are 80% confidence intervals assuming a multivariate t-distribution.

**Table 1.**
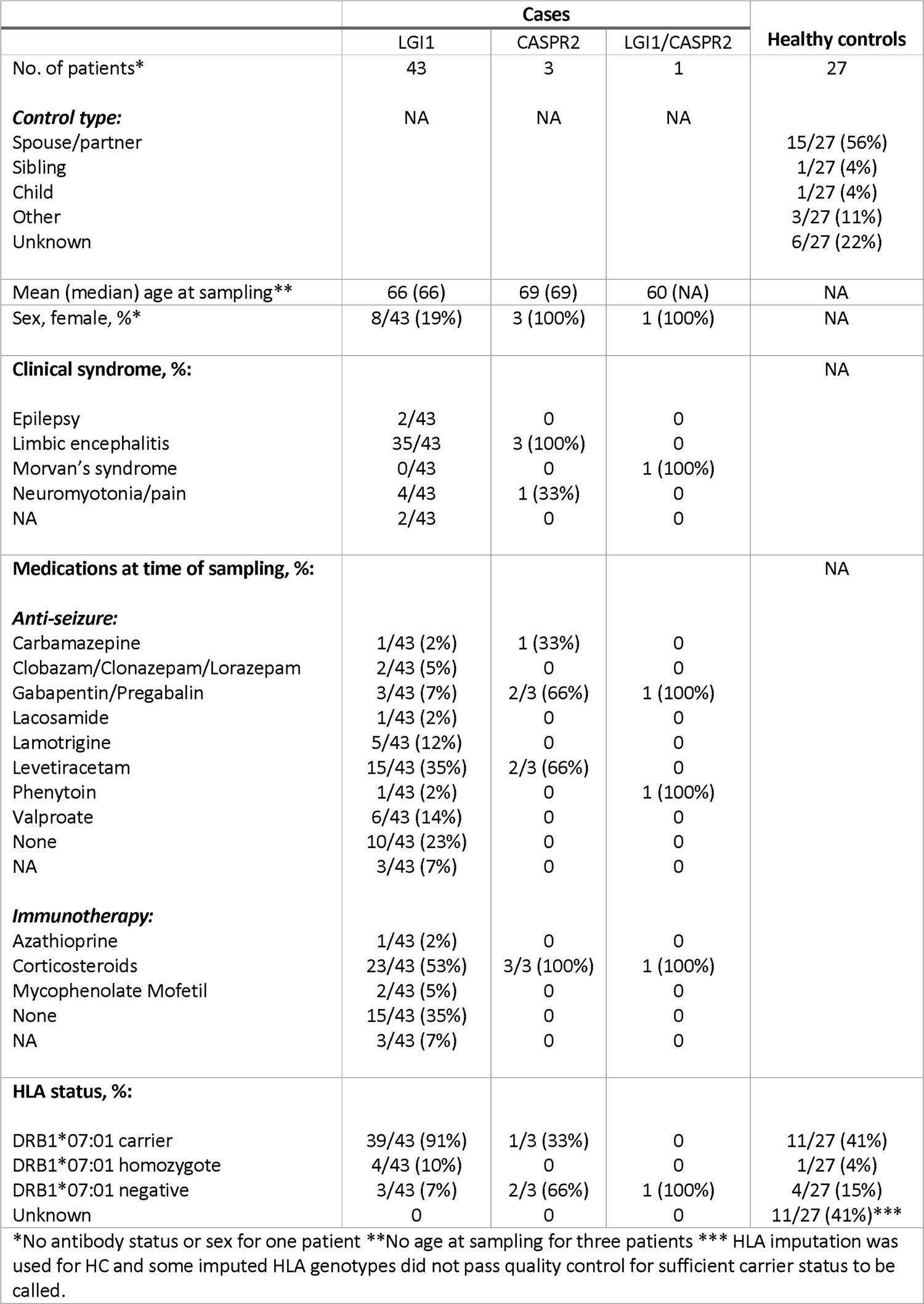
Study Cohort Characteristics. – Shown are the cohort descriptors for 47 LGI1-CASPR2-Ab-E cases and 27 matched health controls. Cohort is characterised by; genetic sex (Sex), auto-antibody status if a case (Case Type), whether an individual is inferred to be a HLA DRB*07:01 carrier through imputation (HLA DRB Status), and what relationship the health control has with matched case (HC Type).

### Gut Microbiome Composition

We first compared the composition of the gut microbiome in cases versus controls. We generated gut microbiome genomic profiles from paired-end metagenomic shotgun sequencing (see Methods and **Supp Figure 2-3**) and investigated composition changes with differential abundance and alpha and beta diversity analysis. The shotgun sequencing generated an average of 8.7 million paired end reads per sample where 84% of these reads passed the adapter trimming and quality filtering stage. An average of 91% of reads passed quality control and were classified using DNA species abundance estimators, *Kraken2* and *Bracken*.

**Figure 2.**
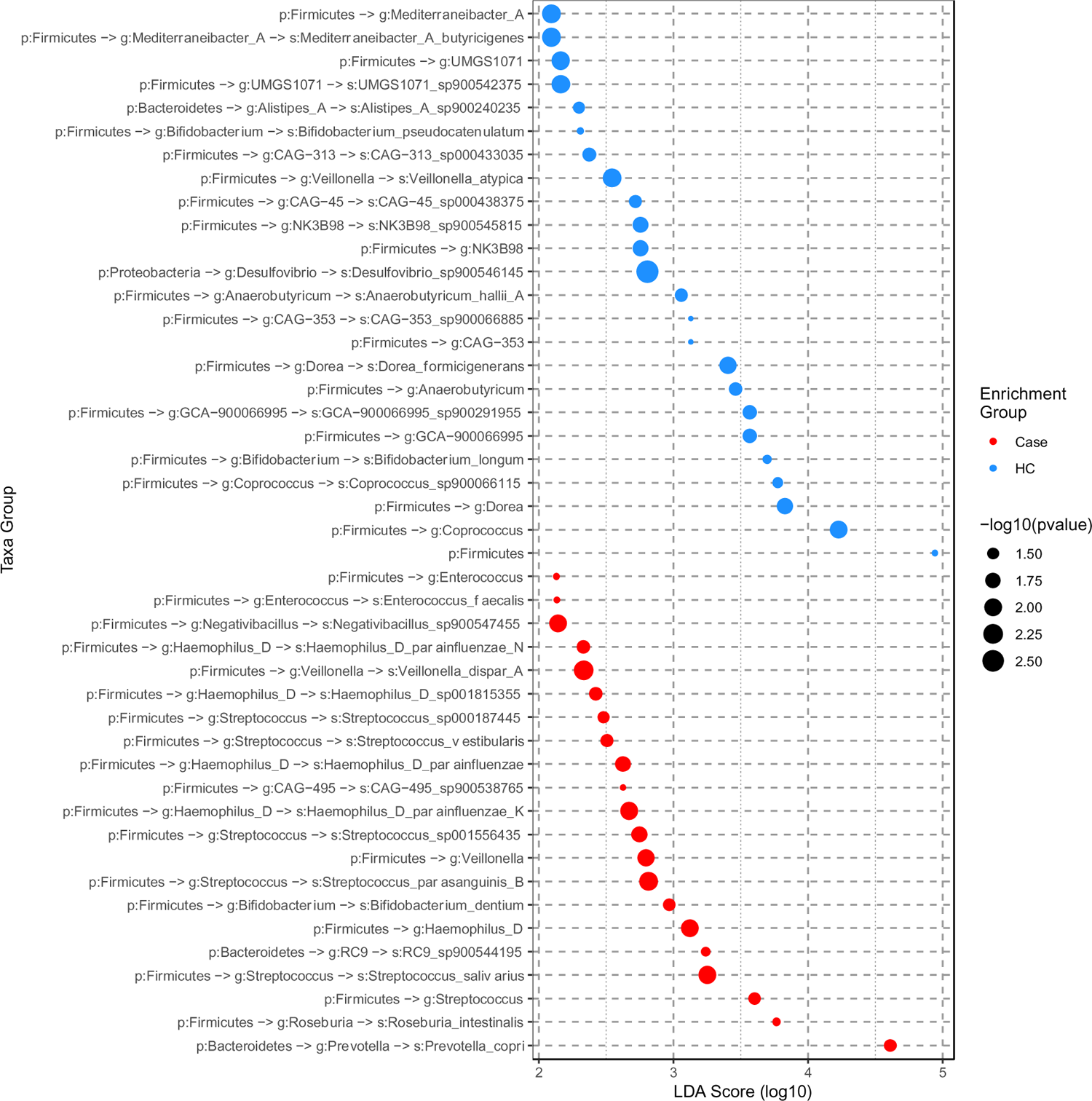
Differential enrichment of microbiome taxa in *LGI1*-*CASPR2*-Ab-E cases and healthy controls-Taxa significantly (adjusted p-value < 0.05) enriched in either healthy controls (HC) or cases. Points are colour coded according to the sample type the taxon is enriched in, and the log_10_( LDA) score (effect size) is shown along the x-axis. Labels along the y-axis are in the format of “p:” to indicate phylum, “g:” to indicate genus, and “s:” to indicate species with “->” joining parent and child taxa.

The general compositional profile of cases and HC were consistent with the expected common human gut microbiome species such as *Bacteroides* or *Faecalibacterium*^29^ (**Figure 1A**). We tested for differences in abundance of individual species and identified no statistically significant differences that survived correction for multiple testing (**Table 2**). Given the rarity of this disease and the fine-grained nature of individual species composition, the 16 most differentially abundant species are reported with uncorrected p-values of < 0.05 (**Table 2**, **Supp Data 2**). Several of the species nominally enriched in LGI1-CASPR2-Ab-E cases are also associated with the human oral and small intestinal microbiome (e.g., *Streptococcus parasanguinis*, *Streptococcus salivarius*, *Bifidobacterium dentium*, and various *Veillonella*).

**Table 2.**
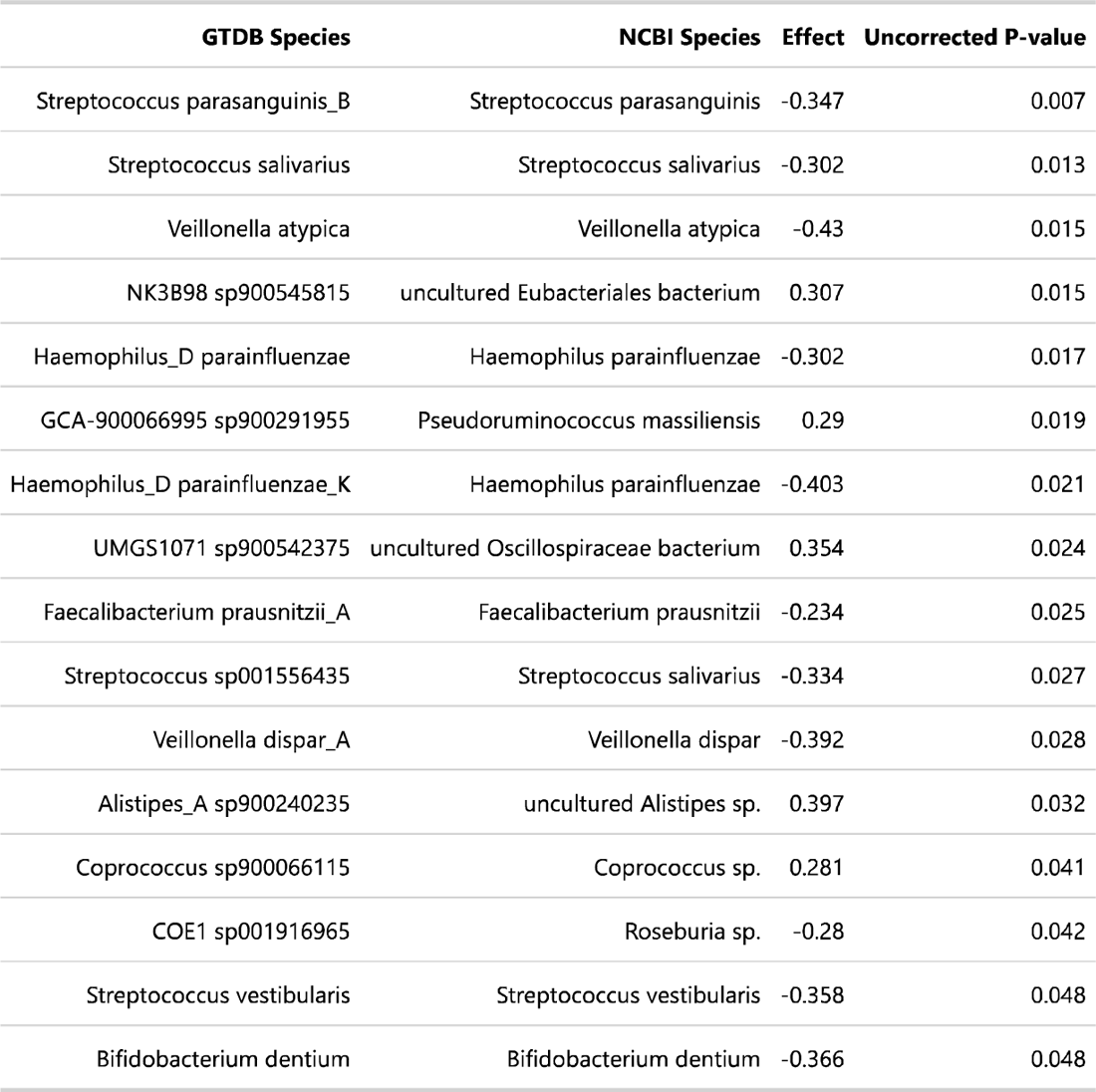
Differential gut microbiome species between *LGI1*-*CASPR2*-Ab-E cases and healthy controls – Shown are the 16 differentiated species which have significant, uncorrected for multiple testing, p-values using the Kruskal-Wallis test using 512 Monte-Carlo instances drawn from the Dirichlet distribution. A positive value indicating an enrichment in HC samples.

To complement and extend this fine-grained species analysis, we simultaneously investigated phyla-, genera-, and species-level enrichment between cases and controls using linear discriminant analysis (LDA) (**Figure 2, Supp Data 3**). Supportive of previous species-level analysis, our LDA results also provide evidence of human oral and small intestinal species of the *Veillonella* and *Streptococcus* genera enriched in cases. In addition, our cases were enriched for the *Enterococcus* and *Haemophilus D* genera, with depletion of the *Dorea* and *Coprococcus* genera, and the *Firmicutes* phylum (recently renamed *Bacillota*^30,31^). Beyond these depletions in HCs, we also observe a depletion specifically of *Bifidobacterium longum* in cases (**Figure 2**).

From the LDA, the differential balance of the *Firmicutes* and *Bacteroidetes* phyla in cases and controls was of interest. Whilst changes in the *Firmicutes/Bacteroidetes* (F/B) ratio has been implicated in some human health-related traits^32–34^, this remains debatable^32^. We investigated such large-scale changes in the context of our modest sample size. As expected^33^ these two Phyla account for the majority of phyla represented in the analysis - 52% + 39.8% (91.2% - Cases) and 30.4% + 61.7% (92.1% - HC). Although the F/B ratio was higher in controls (2.35) than cases (1.21), but this was not statistically significant (Wilcoxon Rank Sum p-value = 0.060) (**Supp Figure 4**).

Considering this F/B ratio difference between cases and controls we explored whether the dosage of HLA-DRB1*07:01 impacts this F/B composition. Analysing the F/B ratio in HC and the LGI1-Ab-E cases (as these were the largest case subtype) (**Supp Figure 4**), we found the F/B ratio to be significantly lower in LGI1 positive cases relative to HCs (cases: 1.21, HCs: 2.35, Wilcox two-sided p-value: 0.032). As expected, we found an enrichment of HLA-DRB1*07:01 heterozygotes in *LGI1*-Ab-E cases versus controls (HWE exact p-value: 3 x 10^-5^) but not in controls (exact p-value: 0.16) (**Supp Figure 5**). We observed the F/B ratio to decrease as a measure of HLA-DRB1*07:01 dosage (Pearson’s product-moment correlation = −0.31 (95% confidence intervals −0.56 to −0.006), p-value = 0.046). The same pattern was no observed in the HCs (p-value = 0.95) (**Supp Figure 5**). Overall, this is the first hint of gut composition differences between *LGI1*-Ab-E cases and controls associated with HLA-DRB1*07:01 genotype.

We next investigated within-gut and between-gut diversity. Testing within-gut with Chao1, Simpson, and Shannon alpha diversity measures we found evidence of subtle differences between case and HC status – consistent with the observations from differential species analysis and LDA. We observed Simpson alpha diversity to be significantly enriched in HC compared to cases (p-value = 0.0313), but not Shannon (Shannon p-value = 0.1112) (**Figure 1B, Supp Figure 6**). Investigating between-gut diversity we performed principal component analysis (PCA) of Aitchison distances of this abundance data. We observed a borderline difference between cases and HC along principal component 2 (p-value = 0.083) (**Figure 1C, Supp Figure 7**). We also observe significant differences between dietary variables and these principal components (**Supp Data 4**), though the uneven sampling size or extremes in scale (such as vitamin most B12 consumption is “infrequent” or “daily”) leads us to caution against these dietary-based observations.

### Functional Characteristics

We next sought to extend the suggestive findings of differences in case-control gut microbiome profiles, characterising function through KEGG modules and pathways^35^ using HUMAnN3^36^ (see Methods). We tested both high-level KEGG modules and lower-level pathways for differential abundance using *Aldex2*^37^, using an uncorrected p-value of 0.05 similar to our differential species analysis (**Table 3, Supp Data 5-6**). We observed three KEGG modules to be significantly different in cases and controls, all enriched in HC. These three modules are particularly interesting in the context of epilepsy. Microbial coenzyme A (CoA) is a key source of short chain fatty acids (SCFA) in the gut^28^, where SCFAs are protective for neuroinflammation and seizure^38^ and epilepsy in general^39^. Ornithine acts as a precursor to CoA in its acetyl-CoA form^40^ and histidine acts as a precursor to ornithine^41^. Of the KEGG pathways all but pentose and glucuronate interconversions (KEGG pathway map00040) were also enriched in HCs. In PCA of these KEGG pathway differences, we observe significant clustering of cases versus HC on principal component three (**Supp Figure 8**) (p-value = 0.044) - though this significant clustering was not found in PCA of KEGG module differences (**Supp Figure 9**).

**Table 3.** Differential KEGG modules between *LGI1*-*CASPR2*-Ab-E cases and healthy controls - Shown are the three differentiated KEGG Modules, and the five KEGG Pathways which have significant, uncorrected for multiple testing, p-values using the Kruskal-Wallis test using 512 Monte-Carlo instances drawn from the Dirichlet distribution. A positive value indicating an enrichment in healthy control samples.

| Table 3 |  |  |  |
| --- | --- | --- | --- |
| Differential KEGG Components |  |  |  |
|  | Name | Effect | Uncorrected P-value |
| **KEGG Module** | Coenzyme A biosynthesis, pantothenate => CoA | 0.468 | 0.0033 |
|  | Ornithine biosynthesis, glutamate => ornithine | 0.369 | 0.023 |
|  | Histidine biosynthesis, PRPP => histidine | 0.314 | 0.037 |
| **KEGG Pathway** | Pentose and glucuronate interconversions | -0.472 | 0.0031 |
|  | Ribosome biogenesis in eukaryotes | 0.358 | 0.0082 |
|  | Parkinson disease | 0.369 | 0.021 |
|  | NOD-like receptor signaling pathway | 0.346 | 0.022 |
|  | Necroptosis | 0.325 | 0.039 |

Finally, we tested the evidence that the gut microbiome of LGI1-Ab-E patients contains potential bacterial homologues of the human *LGI1*, searching the *RefSeq* database for *LGI1* homologues with *blastp* (see Methods for quality cutoffs). We identified 13 organisms to contain such homologues (**Supp Data 7**), however the majority potential homologues are annotated as “leucine-rich” or “hypothetical”, and nearly all organisms in which these homologues are observed are typically found in marine environments. Mapping these potential homologues to the metagenomic data of our LGI1-Ab-E cases, we do not observe any hits and no evidence of their presence in the gut microbiome of LGI1-Ab-E cases.

## Discussion

This metagenomic analysis of the gut microbiome in 47 patients with LGI1/-CASPR2-Ab-E, matched with 27 HC with paired genomic data suggests a potential compositional bias in the gut microbiome of AE. Together our results suggest subtle taxa compositional changes in cases with these differences associated with HLA-DRB1*07:01 dosage. Further this compositional change was associated with depletion of KEGG modules linked to SCFA generation in cases.

We primarily observed that the general gut microbiome of individuals with LGI1/-CASPR2-Ab-E appeared largely consistent with a normal microbiome composition with high abundance of common microbiome species^29^. We therefore infer that LGI1-CASPR2-Ab-E is not characterised by a large difference in microbiome composition driven by specific species. The most positive signal of differing microbiome composition between cases and controls is the enrichment of *Bifidobacterium Longum* in HC. Interestingly, this species has been observed to exert beneficial health effects, on the human gut^42^, inflammation^27^, and potentially reducing cytotoxic effects of certain ASMs^43^.

Instead, linear discriminant analysis highlights several taxa potentially enriched in either our cases or controls, with some of these taxa previously associated with traits relevant to LGI1-CASPR2-Ab-E. Several genera enriched in our cases have been associated with neurological conditions and allows us to place our results into the context of the interplay of the microbiome and neuroinflammation and epilepsy. *Streptococcus*, our strongest signal, has been found to be associated with focal epilepsy in children^44^ where in a small sample (n=10) the authors posit a neuroinflammatory model. Indeed, a mouse model of intestinal inflammation increases convulsant activity and ASM resistance^45^. In addition to *Streptococcus*, the taxa *Enterococcus*^46^ and *Roseburia*^47^ have been found to be associated with intractable childhood or drug-resistant epilepsy (respectively), and *Roseburia* in another study of LGI1-Ab-E^48^. The *genera Coprococcus* and *Dorea* are both enriched in our HC. These have been associated with drug-resistant epilepsy^47^, but *Coprococcus* is also noted for its anti-inflammatory associations^33^ and is depleted in our cases. Our results would support larger, multi-centre studies of the role of the gut microbiome in epilepsy healthcare, from ASM use to disease aetiology and longitudinally from onset to chronic as has been achieved in the cases reported here.

In addition to subtle genera or species differences, our primary observation is the reduction of F/B ratio in LGI1-Ab-E cases compared to HC. This reduction in the F/B ratio appears to be associated with HLA DRB*07:01 allele dosage. The frequency of HLA DRB*07:01 in our cases (91% carriers) matches that observed in cohorts of similar ancestry^19,21^, and our AE cases were primarily LGI1 positive (n=43/47) – in keeping with literature^10^. To our knowledge this is the first observation linking gut microbiome compositional changes to the major LGI1-Ab-E genetic risk locus. Encouragingly a reduction of the F/B ratio and taxa diversity has been associated in another gut microbiome study of LGI1-Ab-E in a cohort of 15 patients of Han Chinese ancestry compared to 25 matched controls using 16S rRNA sequencing^48^. Although the Chinese investigation did not consider HLA genotypes, together our studies show evidence that a subtle shift in taxa composition measured by the F/B ratio is associated with LGI1-Ab-E and is consistent with an inflammatory disease model^48^. That said, for obesity, one of the first traits where the F/B ratio was suggested as a biomarker, results are frequently discrepent^32^ and that phyla encompass a wide variety of taxa with differing functional attributes^26^. Wider and further studies are warranted in neuroinflammatory and epilepsy traits, to investigate the veracity of our observation. These F/B ratio findings appear to be complemented by our functional characteristics results. We find KEGG modules that are nominally depleted in cases, and these modules are associated with CoA synthesis, itself leading to the synthesis of SCFA, which are believed to have an anti-neuroinflammatory effect^39,48^. Taken together with the enrichment of species such as *Bifidobacterium Longum* in HC, our results suggest a model whereby compositional changes in the gut microbiome are associated with functional shifts linked to a loss of neuroprotective SCFA in a context of a specific HLA class II genotype.

In addition to these observed compositional and functional changes, we found in LGI1/-CASPR2-Ab-E cases there is an enrichment of species that are normally associated with the oral or small intestinal microbiomes. It has been observed that corticosteroids impact saliva quality^49^ in a way that could result in accelerated movement of matter though the digestive system, leading to abundance of oral or small intestinal communities. Corticosteroids are a commonly prescribed treatment in of AE^5,6^ as early immune therapy and a majority of our patients have been exposed to this treatment (**Table 1**) - therefore it is a possibility that this observation is driven by treatment and not underlying aetiology. In a linear discriminant analysis comparing cases that have a history of steroid treatment versus case with no history, and versus HCs, we find evidence that this signal is driven by steroid use history (**Supp Figure 10**). Furthermore, there is previous evidence ASMs may also impact the gut microbiota^43^ as does the anti-epileptic ketogenic diet^50,51^. In the latter case, none of the LGI1-CASPR2-Ab-E patients included in this study were or had been on a ketogenic diet.

The primary limitation of this study was cohort size. LGI1-CASPR2-Ab-E is a rare disease which can impact cognitive function, and despite recruitment across the UK and Ireland, only 47 cases were available for analysis. Further, we have studied chronic cases, which may have a different composition to the microbiome of cases with an acute onset. We expect that the sample size constraint impacted our ability to detect more subtle differences between LGI1-CASPR2-Ab-E cases and controls, and limits the statistical power of sub-group analysis. Combining with other AE cohorts of a similar dietary and pharmacological background may provide novel insights, though future care will need to be taken in harmonising gut microbiome sequence generation methods to ensure inter-study comparisons are possible. Compositional changes in the microbiome have been associated with ageing^52^, and we lacked information on the ages of most of our HC. However, we expect our cases and controls to be largely aged-matched (and background matched) as our HC are predominantly spousal or sibling in relationship.

To conclude, our work suggests that LGI1-CASPR2-Ab-E is associated with specific compositional changes in the gut microbiome, shifting the balance between the predominant taxa *Firmicutes* and *Bacteroidetes* in favour of *Bacteroidetes*. This ratio change is associated with the significant HLA risk allelotype DRB*07:01 in cases only and matches previous studies of LGI1-Ab-E cases. This profile is also associated with a shift in functional microbial activity linked to reduced SCFA levels. Given our low sample size, reflective of this rare condition and its effect on statistical power, the exact effect the microbiome plays in disease pathogenesis and/or progression remains unclear. However, our results show a role in a subtle compositional and functional change that is consistent with a neuroinflammatory model and case-specific effect in the HLA DRB*07:01 allelotype. Further work with expanded sample sets and coordinated sequencing protocols appears to be the next step in elucidating the potential trigger for this rare and refractory epilepsy.

## Methods

### Ethics Approval

This study was approved by ethics committees at the Royal College of Surgeons in Ireland (under protocol REC1631), and Oxford University (under protocol number 16/YH/0013). Ethics for this study was approved in the UK by Yorkshire & The Humber - Leeds East Research Ethics Committee (16/YH/0013). All patients gave written informed consent.

### Recruitment

Eligible patients were aged >18 with a diagnosis of autoimmune encephalitis/epilepsy and a positive test for LGI1 and/or CASPR2 antibodies in serum and/or CSF. HCs were spouses, relatives or friends attending clinic or subsequently recruited from the community, preferentially recruiting genetic relatives, or co-habiting relatives. All participants gave informed consent or assent was given by next of kin. Saliva and stool samples were gathered in the home environment and posted via secure biokit. Stool was collected using a Omnigene Gut stool collection kit (DNA Genotek, OM-200). Saliva was collected using Isohelix GeneFix saliva collection kits (Isohelix, GFX-02). Samples were posted to the lab using an overnight tracked-return service and stored at −80°C on arrival. At the time of sampling study participants also completed a questionnaire covering toileting routine, bowel habits, dietary and medication intake (see **Supplementary Data 1** for this questionnaire).

### DNA Extraction

DNA was extracted from stool and saliva samples within two weeks of their arrival at the lab. Saliva extractions were performed using the Isohelix GeneFix Saliva-Prep DNA Kit and stool extractions using the QIAamp PowerFecal Pro DNA Kit (Qiagen, 51804). DNA quality was assessed using a Nanodrop One spectrophotometer.

### SNP Genotyping

Salivary DNA samples were genotyped on the Illumina OmniExpress chip at Edinburgh Genomics, according to manufacturer’s instructions. Genomic ancestry analysis was performed using principal component analysis (PCA) from this SNP genotype data. Case and HC genotypes were merged with Irish and British ancestry controls from the Irish DNA Atlas^53^ (n=193), Irish Trinity Student^54^ (n=2,228), and People of the British Isles^55^ (n=2,039) using PLINK v1.9^56,57^. We filtered for samples and variants with a missingness <5%, and variants with a minor allele frequency <1% and a p-value signifying deviation from Hardy-Weinburg Equilibrium <1e^-9^. We performed PCA on a set of SNPs independent in terms of linkage disequilibrium, using the PLINK --indep-pairwise command with a window of 1000, step size of 50 and linkage filtering threshold of 0.2. In the case of HC, we performed HLA imputation from these SNP genotypes using SNP2HLA^58^

### MB Sequencing

Gut microbiome DNA samples were shipped to SeqBiome Ltd, Fermoy, County Cork, Ireland. Library construction and shotgun sequencing of autoimmune cases and controls were carried out at SeqBiome Ltd using Nextera library preparation^59^ and Illumina NovaSeq sequencing (2 × 150 bp per sample, Illumina, Hayward, California, USA).

Gut microbiome sequence data was then processed initially with *FastQC*^60^, where data quality was visually inspected. All samples were of generally good quality with minimal quality drop off at the end of sequences. *Trimmomatic*^61^ was used for trimming and quality filtering using the following parameters: SLIDINGWINDOW:5:22, MINLENGTH:75. The data was then passed to *Kneaddata*, a pipeline incorporating *Trimmomatic*, *Bowtie 2* ^62^, and other elements designed for contaminant removal. Taxonomic assignment was then performed using *Kraken2* using a confidence of 0.1 and the *Kraken2*^63,64^ GTDB bacterial database^65^. *Kraken2* is a taxonomic classification system using exact k-mer matches to achieve high accuracy and fast classification speeds, using a customised version of the GTDB database, the results of which were analysed using the R based phyloseq package^66^. GTDB is a database which clusters available genomes based on Average Nucleotide Identity (ANI), and assigns taxonomy based on the National Center for Biotechnology Information (NCBI) classifications. If a sequence does not meet clustering requirements (95% ANI), but does not have a unique taxonomic classification, a suffix will be added to indicate this (e.g., *Escherichia coli* A). This allows for the most accurate taxonomic classifications, even in the case of organisms which have yet to be formally identified.

Pathway and gene family assignment was then performed using *humann3*^36^. The standard Uniref90 annotation of gene families were collapsed into Kyoto Encyclopedia of Gene and Genomes (KEGG)^35^ orthology annotation and further collapsed into KEGG module and then pathway annotation.

### Comparative Analysis

We used the phyloseq package^66^ in R^67^ v4.1.3 for comparative alpha diversity, beta diversity and taxonomic analysis, using the package *ALDEx2*^37^, to determine if any species were differentially abundant (at group level) based on a Kruskal-Wallis test, using 512 Monte-Carlo instances drawn from the Dirichlet distribution. P-values were initially corrected for Benjamini-Hochberg multiple testing, and all were > 0.05. We therefore focussed discussion on the species with P-values significant prior to multiple testing correction (< 0.05 prior to correction).

To account for the compositional structure of the Next Generation Sequencing (NGS) generated data and to avoid the likelihood of generating spurious correlations, we first imputed the zeros in the abundance matrices using the count zero multiplicative replacement method (cmultRepl, methodlll=lll”CZM”) implemented in the *Compositions* package and applied a centred log-ratio transformation (CLR) using the *codaSeq.clr* function in the *CoDaSeq* package.

Linear discriminant analysis (LDA) effect size estimation was carried out using the *run_lefse()* function *microbiomeMarker* package in Bioconductor on the species abundance matrix, selecting CPM normalisation method, 100 bootstrap replicates, a Kruskal-Wallis and Wilcoxon p-value cutoff of 0.05 and LDA effect size cutoff of 2. To test HLA-DRB1*07:01 divergence from Hardy-Weinberg-Equilibrium (HWE) expectations we utilised the R package HardyWeinberg and the function HWExact() to calculate an exact test for the HWE. To test correlations between HLA dosage and F/B ratio we used the R function cor.test().

Alpha and Beta diversity analysis was carried out using the *R* packages *vegan* and *phyloseq* and statistical comparison was tested using Kruskal-Wallis tests. Principle Coordinate Analysis (PCA) was carried out using the *PCA* function in *R* using Aitchison distance matrix (CLR + Euclidean distances). To test the significance of groupings in PCA space, Permutational Multivariate Analysis of Variance (PERMANOVA) test was then performed using 999 permutations with significance cut-off value of 0.05. Dispersion homogeneity was tested using the *betadisper* function from *vegan*.

### Functional Comparison

KEGG pathways and modules were assessed for differential abundance using *Aldex2* based on a Kruskal-Wallis test using 512 Monte-Carlo instances drawn from the Dirichlet distribution. P-values were initially corrected for multiple testing, and all were > 0.05. We therefore focussed discussion on the pathways and modules with P-values significant prior to multiple testing correction (< 0.05 prior to correction). KEGG pathways and KEGG modules were analysed by computing PCAs using Aitchison distances. To test the significance of groupings, PERMANOVA tests were performed on these distances, using 999 permutations, with a p-value cut-off of 0.05. Dispersion homogeneity was tested using the *betadisper* function from *vegan*.

To assess the presence of potential bacterial homologs of the human *LGI1* protein, a *blastp* search of the *LGI1* protein was run using a database of 64,695 bacterial genomes acquired from the *RefSeq* database from which proteomes were generated using *Prokka*^68^. Potential homologs were then filtered from these results using cutoffs of >40% identity, >10% coverage and an e-value <1e-10. These potential homologs were then mapped back to the metagenomic data using *blastx* with cutoffs of >80% identity, >80% coverage and an e-value of <1e-6.

## Data Availability

The raw metagenomic sequence data will be made accessible on the European Genome-Phenome Archive (EGA) upon publication and will be referred to here.

## Acknowledgements

We would like to thank all the participants of this study, without whom the work could not have been completed. We would like to thank the help of Bullers and Mrs Rachel Feeney from the OCMS for their help with kit assembly and sample extractions.

## Open access

For the purpose of Open Access, the author has applied a CC BY public copyright licence to any Author Accepted Manuscript (AAM) version arising from this submission.

## Supplementary Materials

**Supplementary Figure 1.**
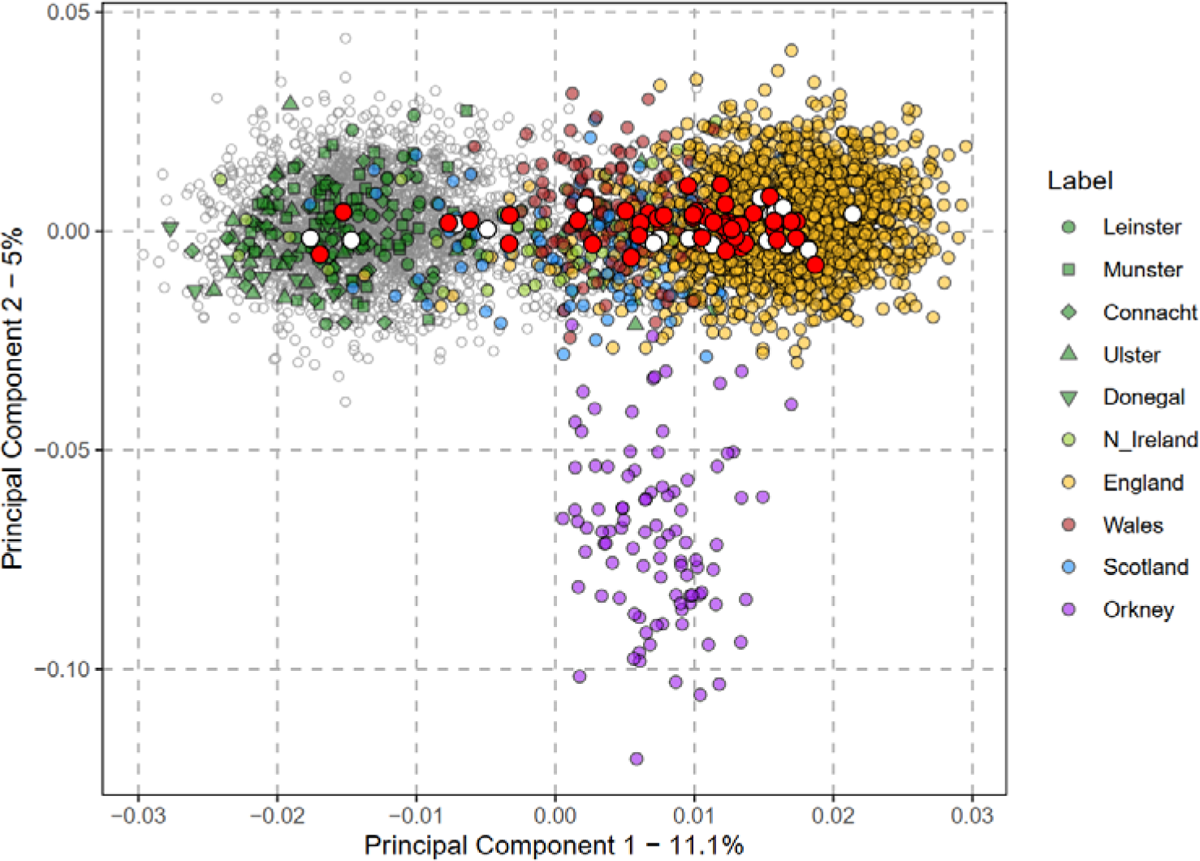
Ancestry PCA of *LGI1*-*CASPR2*-Ab-E Cases and Controls. . The principal component decomposition of a genetic relationship matrix of 4,460 Irish and British ancestry controls and 47 cases and 27 health controls (HC). Points are colour and shape coded according to genetic ancestry group. Filled circles with red indicate the genetic position of a case and filled circles with white indicate the genetic position of a HC.

**Supplemental Figure 2.**
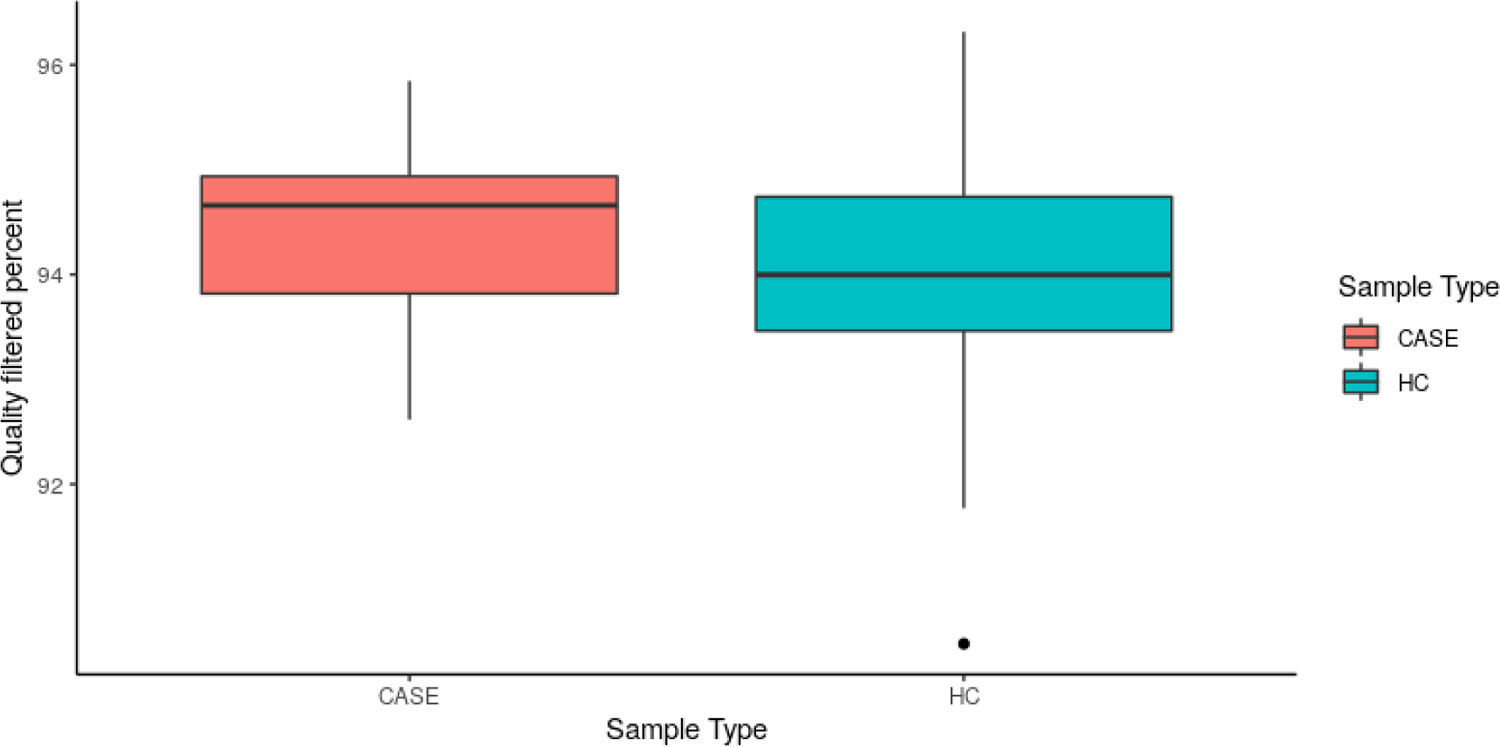
Filtering quality of *LGI1*-*CASPR2*-Ab-E case and healthy control gut microbiome samples. Boxplot representation of percentage of reads which pass the quality filtering step, coloured by sample type. Boxplots show the median value, with lower and upper hinges showing the 1st and 3rd quartiles. Whiskers show the largest value no further than 1.5 x the Interquartile Range (IQR) from that range. Data points beyond these whiskeys are plotted separately as black points.

**Supplemental Figure 3:**
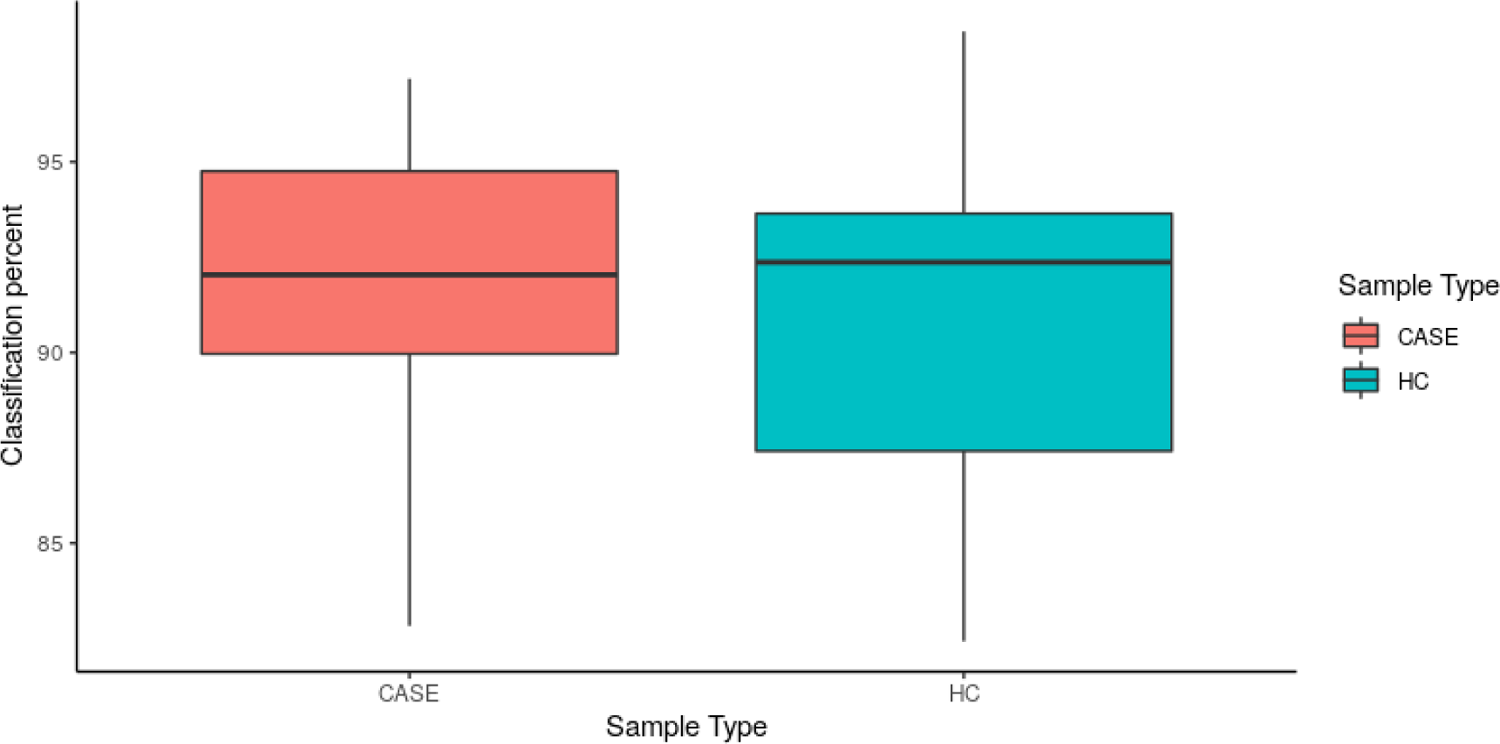
Kraken2 percent classification of *LGI1*-*CASPR2*-Ab-E case and healthy control reads. Boxplots show the median value, with lower and upper hinges showing the 1st and 3rd quartiles. Whiskers show the largest value no further than 1.5 x the Interquartile Range (IQR) from that range.

**Supplemental Figure 4:**
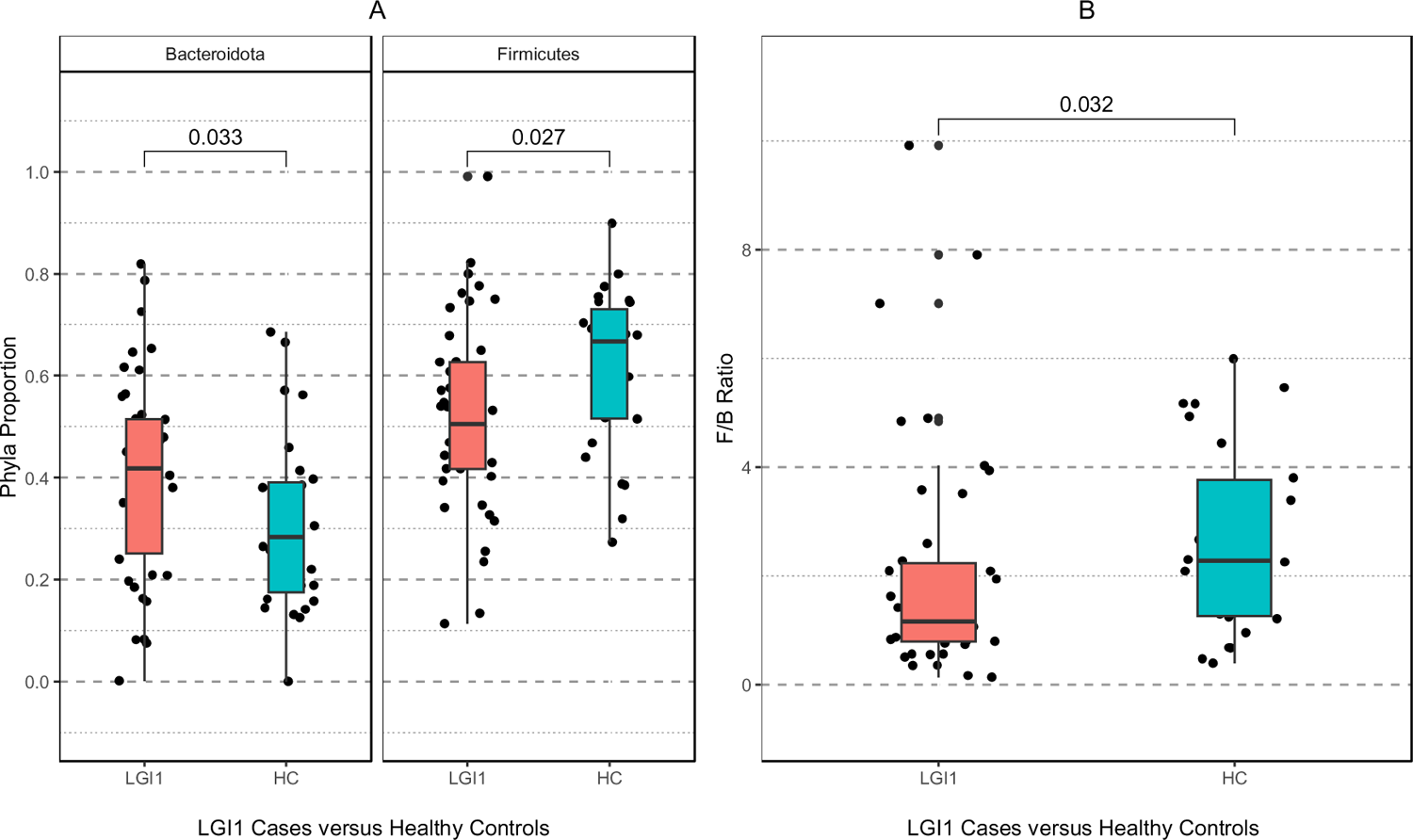
*Firmicutes*/*Bacteroides* ratio in *LGI1*-*CASPR2*-Ab-E cases and healthy controls. (A) The proportion of the two phyla *Firmicutes* and *Bacteroides* in cases and healthy controls (HC). Comparison bars shown are the p value from Wilcox Signed Rank test. (B) The *Firmicutes*/*Bacteroides* ratio between cases and HC. Individual data points are shown on top of boxplots. Boxplots show the median value, with lower and upper hinges showing the 1st and 3rd quartiles. Whiskers show the largest value no further than 1.5 x the Interquartile Range (IQR) from that range.

**Supplemental Figure 5:**
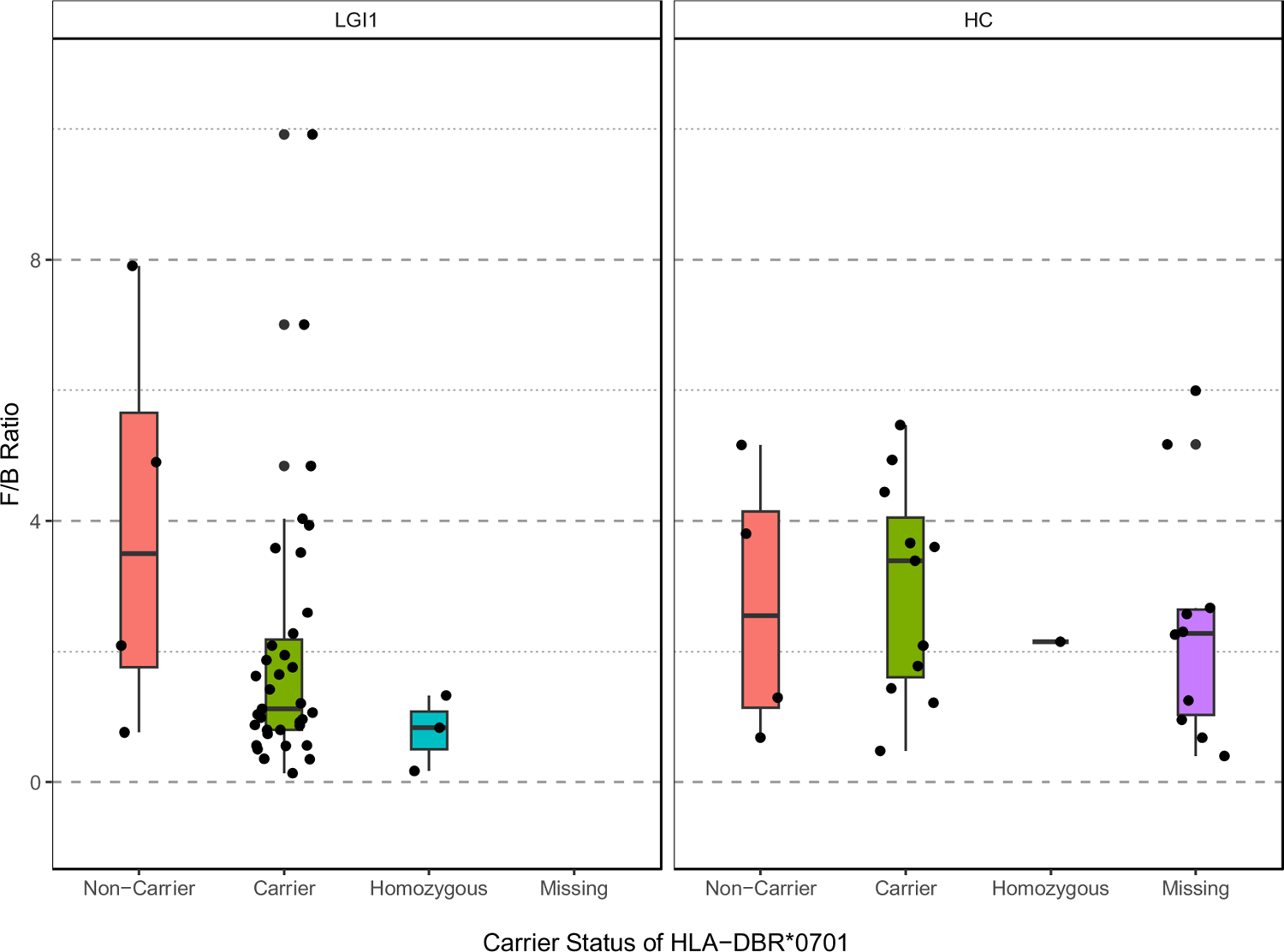
*Firmicutes*/*Bacteroides* ratio as a function of HLA-DBR*0701 dosage. The ratio of the two phyla *Firmicutes* and *Bacteroides* in LGI1-Ab-E cases and healthy controls (HC) grouped by HLA-DBR*0701 dosage. As HC HLA-DRB1*0701 dosage was imputed from SNP genotypes there is a proportion of missing data. Individual data points are shown on top of boxplots. Boxplots show the median value, with lower and upper hinges showing the 1st and 3rd quartiles. Whiskers show the largest value no further than 1.5 x the Interquartile Range (IQR) from that range.

**Supplemental Figure 6.**
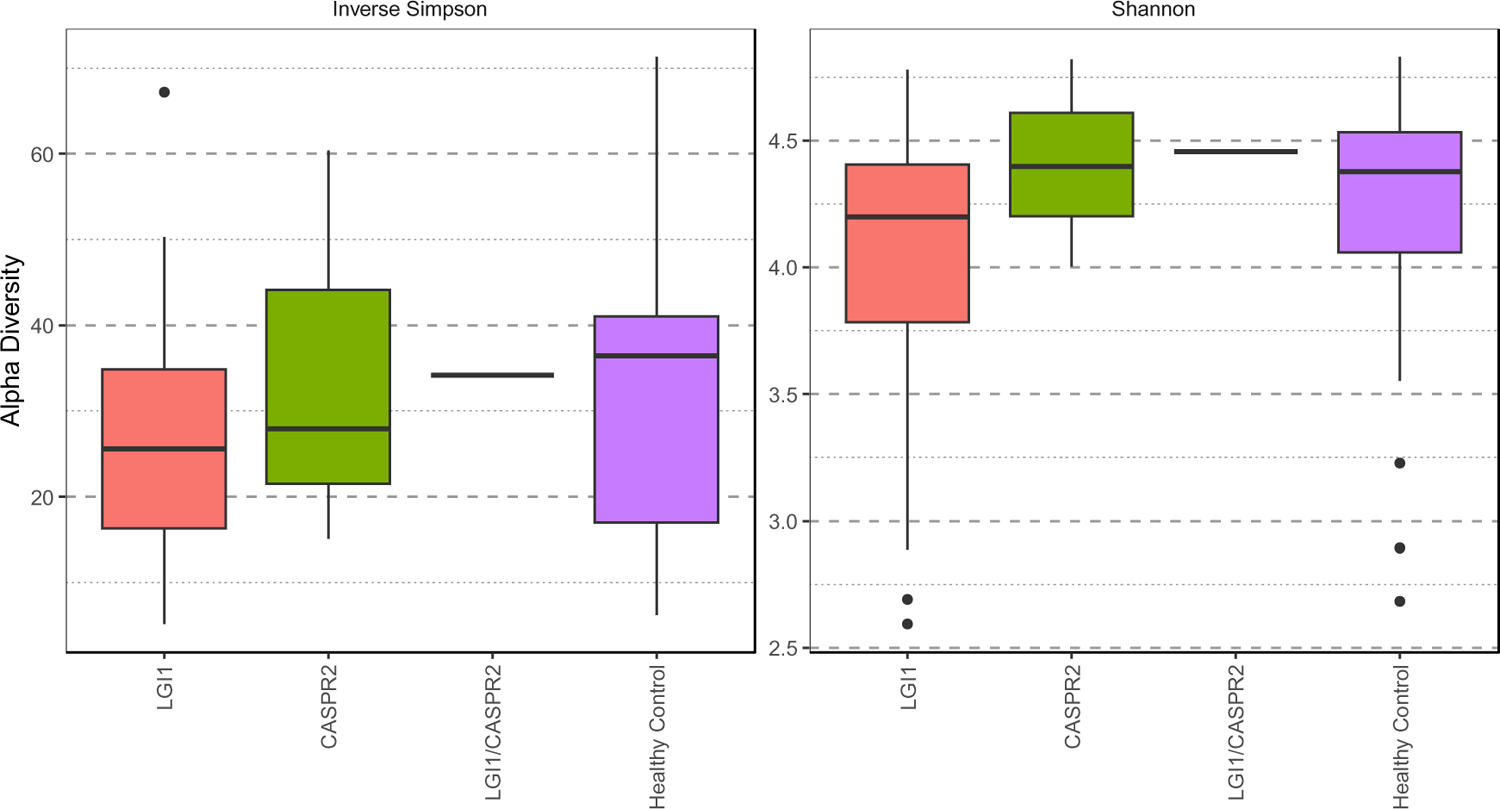
Measure of Alpha diversity stratified by case type. Shown are the Inverse Simpson and Shannon alpha diversity measurements. Distributions of alpha diversity values are shown in boxplots. Boxplots show the median value, with lower and upper hinges showing the 1^st^ and 3^rd^ quartiles. Whiskers show the largest value no further than 1.5 x the Interquartile Range (IQR) from that range. Data points beyond these whiskeys are plotted separately as black points.

**Supplemental Figure 7.**
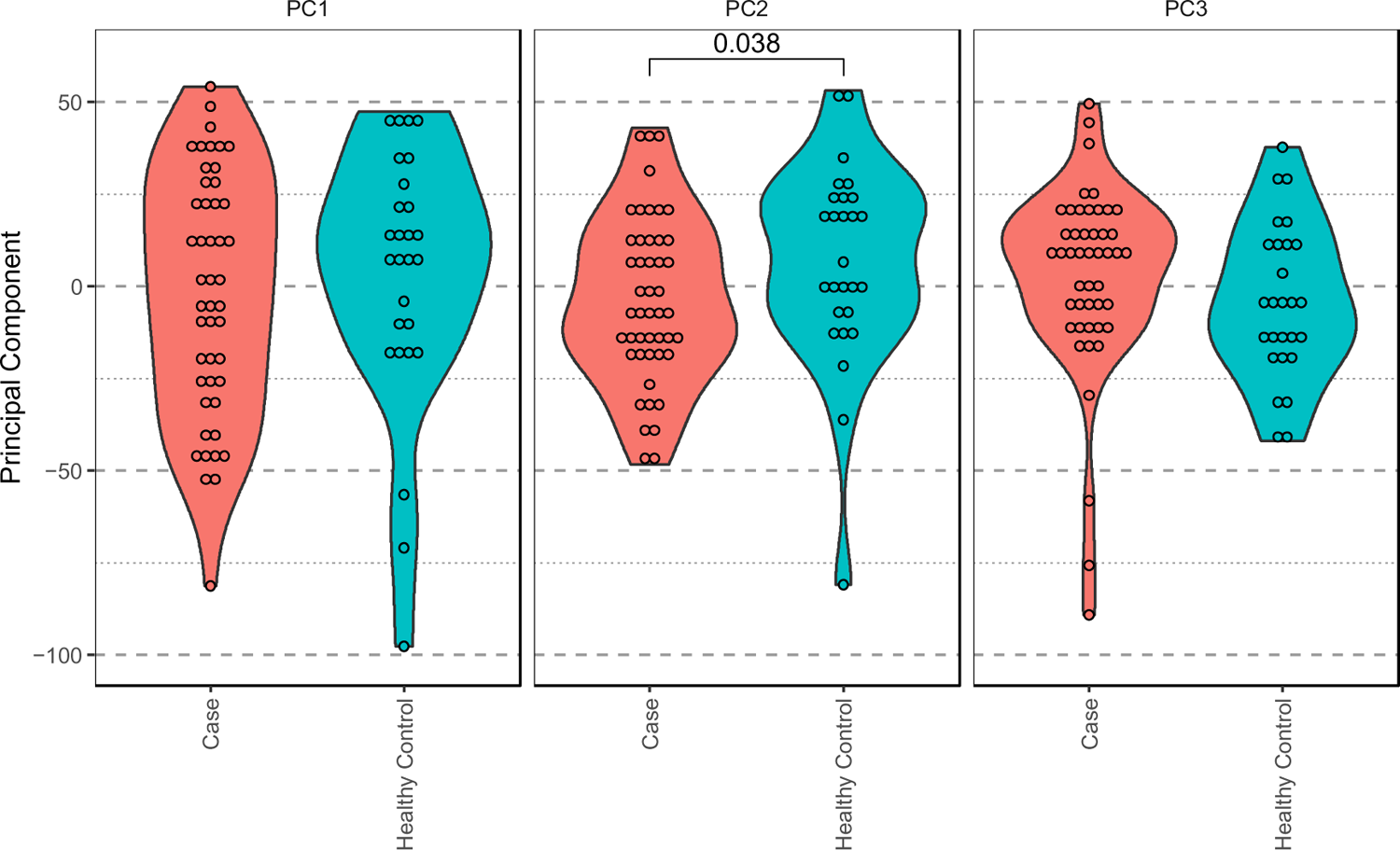
Distribution of LGI1-CASPR2-Ab-E cases and controls over beta-diversity principal components. Shown are the sample distributions across the first three Aitchison principal components of beta diversity, with violin plots showing overall distribution. Pairwise comparison values are significant p-values from Wilcox tests of significant differences. All comparison pairs were tested, and p-values adjusted with the Holm method.

**Supplemental Figure 8.**
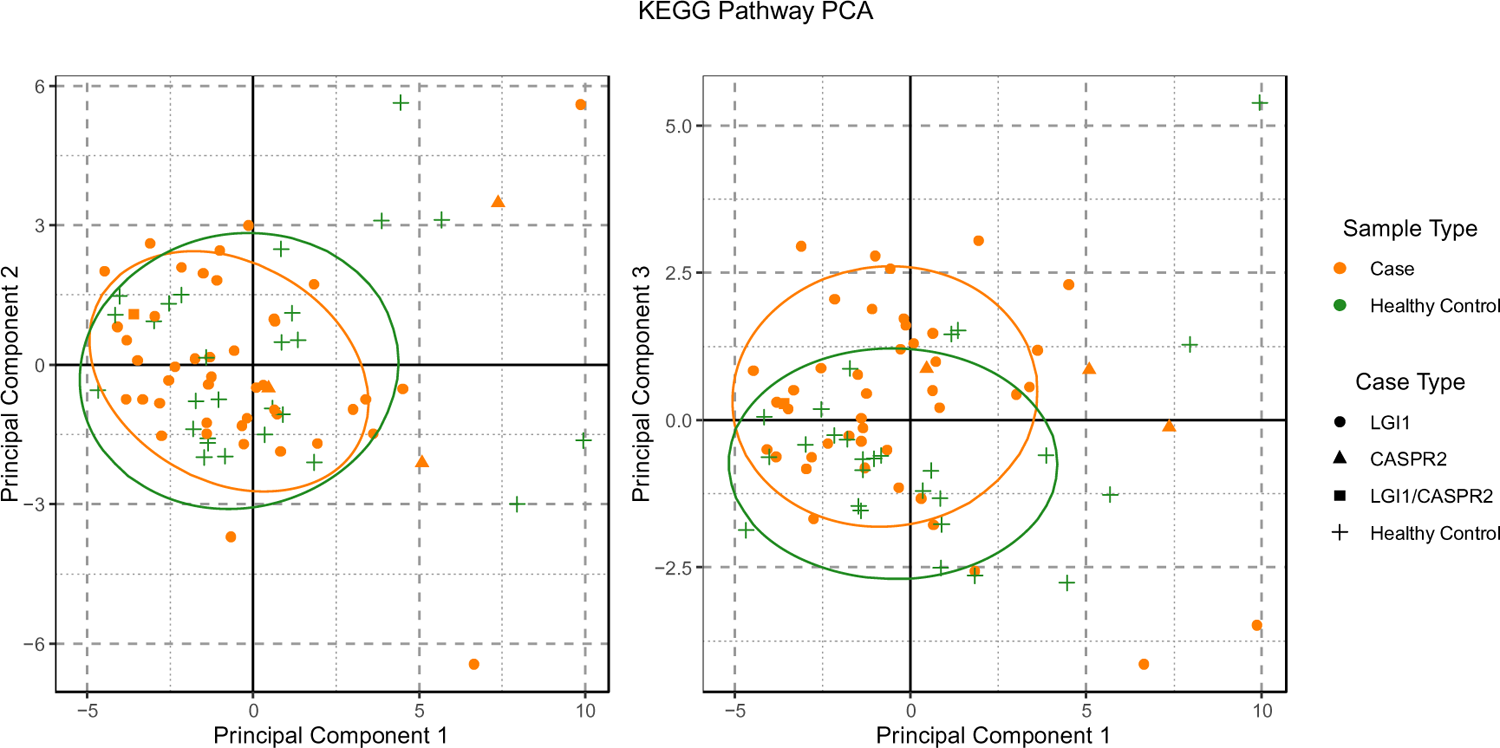
Aitchison PCA of KEGG Pathway abundance. Sample case and control type are shown by colour and shape. Ellipses shown are 80% confidence intervals assuming a multivariate t-distribution.

**Supplemental Figure 9.**
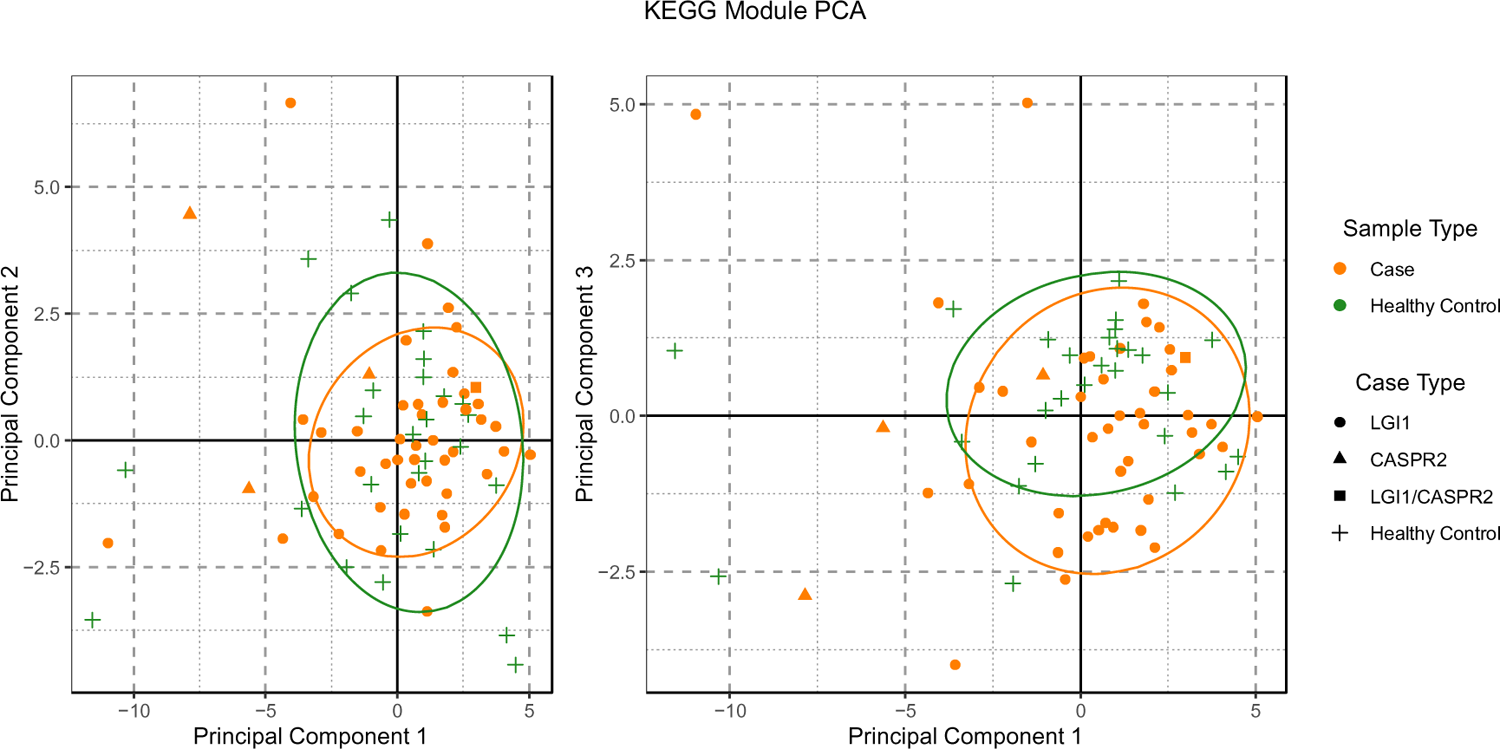
Aitchison PCA of KEGG Module abundance. Sample case and control type are shown by colour and shape. Ellipses shown are 80% confidence intervals assuming a multivariate t-distribution.

**Supplemental Figure 10.**
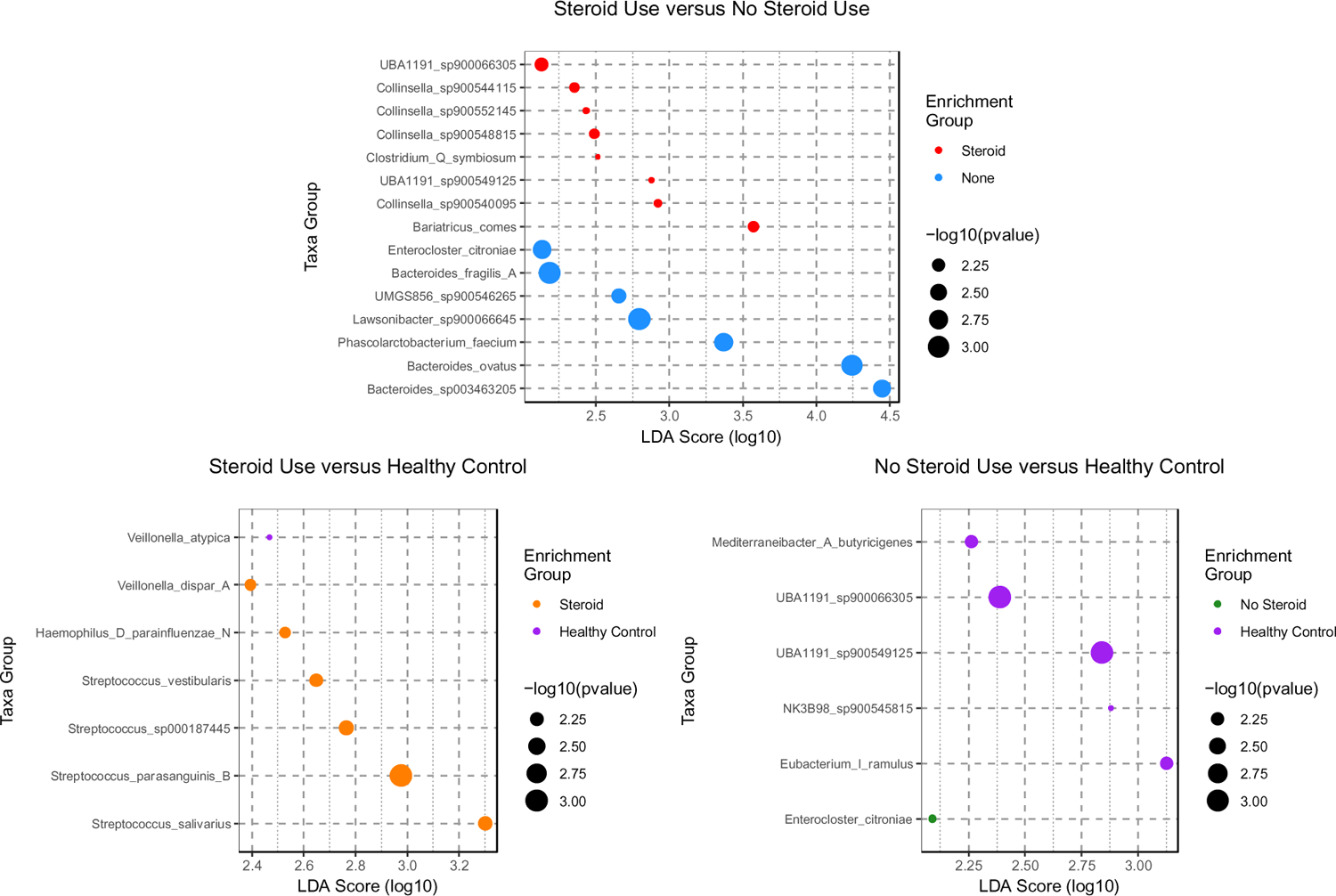
Differential enrichment of microbiome taxa in *LGI1*-*CASPR2*-Ab-E cases stratified by steroid use history and healthy controls-Taxa significantly (adjusted p-value < 0.05) enriched in either healthy controls (HC) or cases. Points are colour coded according to the sample type the taxon is enriched in, and the log_10_( LDA) score (effect size) is shown along the x-axis. Labels along the y-axis are in the format of “p:” to indicate phylum, “g:” to indicate genus, and “s:” to indicate species with “->” joining parent and child taxa.

**Supplemental Data 1 – Dietary and supplement questionnaire**

**Supplemental Data 2 – Species**

**Supplemental Data 3 – LDA Taxa results**

**Supplemental Data 4 – Dietary associations with beta-PCs**

**Supplemental Data 5 – All KEGG Module results**

**Supplemental Data 6 – All KEGG Pathway results**

**Supplemental Data 7 – Organism orthologues**

